# Assessment of safe wheeled walker use in frail older adults: Development of a video-based rating instrument

**DOI:** 10.64898/2026.06.04.26354904

**Authors:** Rebekka Leonhardt, Ulrich Lindemann, Marc Schneider, Kilian Rapp, Jochen Klenk

## Abstract

**Background:** Wheeled walkers can improve safety during walking, but improper use may increase fall risk among frail older adults. No suitable tool exists to assess safe indoor wheeled walker use in this population. This study aimed to develop and validate a video-based expert assessment tool.

**Methods:** Based on the literature and expert consensus, seven problematic indoor situations were identified, and an assessment tool with five safety criteria per situation was developed (maximum score = 35). Fifty participants (mean age 83.9 years, 64% women) from a geriatric rehabilitation clinic and a nursing home were video-recorded while using a rollator. Expert ratings were compared with nursing staff ratings, self-ratings, and the Timed Up and Go test to evaluate validity. Intra- and inter-rater reliability were determined from independent ratings by two physiotherapists and a repeated expert rating after seven days. Sensitivity to change was assessed after two weeks of rehabilitation, and feasibility by the time required for assessment.

**Results:** The expert score of rater 1 at baseline was 28.5 points, and assessment required a mean of 17.5 minutes. Intra-rater reliability was excellent (ICC = 0.98) and inter-rater reliability was good (ICC = 0.80). Validity analyses showed the strongest association with nursing staff assessments (r = 0.74) and a moderate association with the Timed Up and Go test (r = –0.45). After two weeks, patients improved by an average of 2.38 points (8.4% of baseline score).

**Conclusions:** The new instrument demonstrated high reliability, acceptable validity, sensitivity to change, and good feasibility for assessing safe wheeled walker use in frail older adults.

**Trial registration number and date of registration:** DRKS00038358, 07/11/2025

## Introduction

Frail older adults, such as geriatric patients or nursing home residents, experience one to two falls per person-year, with most falls occurring during sit-to-stand transfers and walking [1].

The causes of transfer and gait instability are manifold, including muscle weakness and balance deficits. In addition to the personal consequences for those who fall (fractures, bruises, sprains, anxiety, social withdrawal), falls lead to substantial healthcare costs and an increased workload for nursing staff. To compensate for age-related declines in strength and balance, wheeled walkers are often used to stabilize gait and increase mobility and participation [2]. Previous studies have demonstrated improvements in walking performance [3, 4], reductions in the severity of fall-related injuries [5], and decreased fear of falling [6] associated with wheeled walker use.

However, wheeled walkers may themselves become a source of risk if used improperly. The increased fall risk associated with wheeled walker use [7] may be partly explained by an interaction between users’ underlying gait instability and improper handling of the device in specific contexts. Hazardous indoor situations, such as opening doors or walking backwards while using a wheeled walker, have been identified in a survey of wheeled walker users [8].

One existing tool designed to evaluate wheeled walker safety in people with dementia [9] includes outdoor problem situations, limiting its applicability in nursing homes and clinical settings. Furthermore, this instrument does not weight problem situations uniformly and incorporates assessments of physical performance that are not necessarily related to safe wheeled walker use. An assessment instrument capable to identifying individual, situation-specific problems encountered by frail older adults using a wheeled walker, which would be essential for guiding targeted therapeutic interventions, is currently not available. To address this gap, the aim of the present study was to develop an instrument for evaluating the safe indoor use of wheeled walkers in frail older adults.

## Materials and Methods

### Subjects and design

This development study has a cross-sectional part (validity, reliability) and a prospective part (sensitivity to change). Participants were recruited from a geriatric rehabilitation clinic and a nursing home in south-western Germany between July 15, 2024, and April 17, 2025. During the recruitment phase, the study setting was expanded to additionally include nursing home residents in order to evaluate the feasibility of the assessment in this setting. This protocol amendment did not affect the assessment procedures. Inclusion criteria were an age of 60 years or older and independent use of a four-wheeled walker (rollator). Exclusion criteria were cognitive impairment (as rated by the responsible physician in the hospital or by nursing staff in the nursing home), insufficient German language skills, and terminal illness. The study protocol was approved by the Ethics Committee of the State Chamber of Physicians of Baden-Württemberg, Germany (F-2024-038). All participants provided written informed consent.

### Baseline characteristics of the participants

Descriptive data, including age, body mass index, and medical diagnoses, were extracted from the electronic health records of the hospital and nursing home during the above-mentioned recruitment period. Prior experience with wheeled walker use and fall history (preceding 12 months) were obtained by interview. Performance on the Timed Up and Go (TUG) test [10] was measured by stopwatch from the video-recorded assessment.

### Development of the test protocol

In the first step, two focus group meetings were conducted with four physiotherapists experienced in geriatric care. Based on the literature and the experts’ clinical experience, nine potentially problematic situations related to indoor wheeled walker use were identified. For each situation, five safety criteria were defined (e.g. no collision of feet with wheeled walker, adequate brake handling, sufficient distance between feet and wheeled walker). In the second step, the protocol was pilot-tested with two patients, after which two situations were removed due to overlap with others. In the third step, the protocol was tested with two additional patients and finally approved by the study team.

### Test protocol

Participants were video-recorded while performing seven potentially problematic indoor situations: (1) standing up from a chair, (2) walking 3 m around a cone and back, (3) sitting down again, (4) opening a door in the direction of walking and passing through it, (5) turning on the spot (180°), (6) opening a door against the direction of walking and passing through it, and (7) sitting down on the wheeled walker and standing up again. Three videos were created per participant: the first included situations 1-3, the second situations 4-6, and the third situation 7. In the geriatric rehabilitation setting, the video recordings were made at the start of the rehabilitation phase on the clinic’s premises. In the nursing home setting, the video recordings were made in the immediate vicinity of the residents’ rooms.

Using the video recordings, a physiotherapist rated each of the seven situations according to five predefined safety criteria, yielding a maximum score of 35 points, with higher scores indicating greater safety. The full protocol, including all problematic situations and safety criteria, is provided in the **S1 Appendix**. The time frame for the baseline assessment was set for 30-45 minutes.

For validity evaluation, participants rated their perceived safety during each situation on an 11-point Likert-scale (0-10). In addition, the nurse in charge rated overall safety based on the video recordings using the same scale. Both scores were rescaled to 0-35 points to improve comparability with the expert ratings. The time required to complete the TUG test, derived from the video recordings, was also used for validity evaluation.

Reliability of the video ratings was assessed by repeated ratings of the same videos by the same physiotherapist after seven days (intra-rater reliability) and by a second physiotherapist (inter-rater reliability). Sensitivity to change was assessed by repeating the video-based assessment after two weeks of standard rehabilitation. Feasibility of the tool was evaluated by recording the time required for video recording and rating.

### Statistics

Statistical analyses were conducted in R (version 4.5.3; R Core Team, 2026). Descriptive characteristics are presented as means with standard deviations (SD) or confidence intervals (CI), minimum and maximum values, or counts (n) and percentages (%), as appropriate.

Validity was examined by calculating associations between measures using Pearson’s correlation coefficient (r). Reliability was assessed using intra-class correlation coefficients (ICCs) for inter-rater (two raters) and intra-rater (same rater at baseline and seven days) agreement. Sensitivity to change was analysed by calculating the mean difference between baseline and two-week follow-up scores in patients undergoing standard rehabilitation that did not specifically focus on wheeled walker use.

## Results

Data from 50 participants (geriatric rehabilitation clinic: n = 40; nursing home: n = 10) including 32 women (64%), with a mean age of 83.9 years were available for analysis. **Figure 1** illustrates participant’s progression through enrolment, baseline assessment, follow-up assessment, and data analysis.

**Figure 1.**
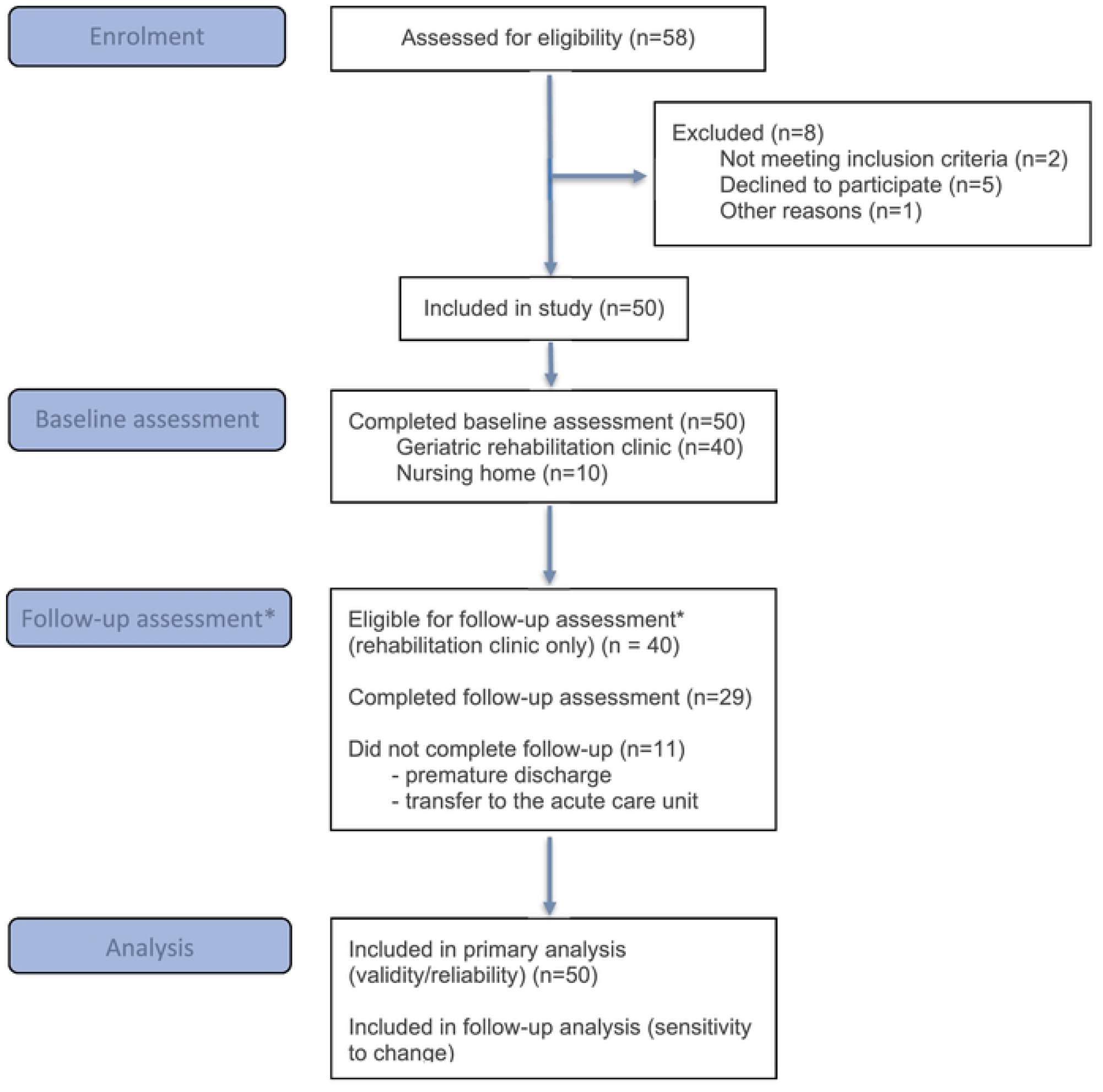
CONSORT Flowchart of participant progression through enrolment, baseline assessment, follow-up assessment, and data analysis. ^*^Follow-up assessment were only feasible in participants recruited in the geriatric rehabilitation clinic setting. Citation: Hopewell S, Chan AW,Collins GS, Hr6bjartsson A, Moher D, SchulzKF,et al. CONSORT2025 Statement: updated guideline for reporting randomised trials. BMJ.2025; 388:eOS1123. https://dx.doi.org/10.1136/bmj-2024-081123 © 2025Hopewell et al. This is an OpenAccessarticle distributed under the terms of the Creative Commons Attribution License(https://creativecommons.org/licenses/by/4.0/). whichpermits unrestricted use,distribution, and reproduction in any medium, provided the original work is properly cited.

Mean performance in the TUG test was 34.9 seconds, with all participants performing worse than 15 seconds. Baseline characteristics of the cohort are presented in **Table 1**. No adverse events occurred during the assessments.

**Table 1.** Baseline characteristics of all 50 participants including 40 patients of a geriatric rehabilitation clinic and 10 nursing home residents.

|  |  |  |
| --- | --- | --- |
| Sex, n (%) | Female | 32 (64) |
|  | Male | 18 (36) |
| Age [years], mean (SD, min-max) |  | 83.90 (7.07, 65.00–98.00) |
| Body mass index [kg/m<sup>2</sup>], mean (SD, min-max) |  | 25.75 (6.44, 16.75–55.77) |
| Timed-Up-and-Go [s], mean (SD, min-max) |  | 34.94 (11.95, 16.09–64.80) |
| Fall history (last 12 months), n (%) | no | 28 (56) |
|  | yes | 22 (44) |
| Previous wheeled walker experience, n (%) | no | 23 (46) |
|  | yes | 27 (54) |
| Main diagnoses, n (%) | fracture | 19 (38) |
|  | other orthopaedic problem | 4 (8) |
|  | neurologic disease | 12 (24) |
|  | Deconditioned | 15 (30) |

The mean safe wheeled walker use scores assigned by Rater 1 and 2 at the initial assessment were 28.5 and 30.1 points, respectively, with only one participant achieving the maximum score of 35 (**Table 2, Figure 2**). Safety criteria rated as negative in more than 30% of all ratings identified brake handling, turning on the spot (180°), sitting down on the walker, and opening a door as the most relevant problem areas. Participant self-ratings and staff ratings were lower than expert ratings. Mean assessment duration, including video recording and rating, was 17.5 minutes.

**Table 2.** Rating results of experts, participants and nursing staff, as well as measurement duration.

|  |  |  |
| --- | --- | --- |
| Expert rating of safe wheeled walker use [0-35], mean (SD, min-max) | Rater 1, 1<sup>st</sup> rating | 28.5 (4.67, 13-35) |
|  | Rater 1, 2<sup>nd</sup><br>rating after 7d | 28.7 (4.72, 13-35) |
|  | Rater 2 | 30.1 (4.22, 17-35) |
| Participants self-rating of their safety during<br>each situation [0-35]*, mean (SD, min-max) |  | 25.3 (5.90, 11-35) |
| Nursing staff rating of the overall safety<br>based on the video recordings [0-35]*, mean<br>(SD, min-max) |  | 22.9 (8.22, 7-35) |
| Measurement duration [min] (including video<br>recording and rating, mean (SD, min-max) |  | 17.47 (3.79, 12.38–29.50) |

\* Self-rating (0-70) and nursing staff-rating (0-10) were rescaled to 0-35 points to improve comparability between the safety scores.

**Figure 2.**
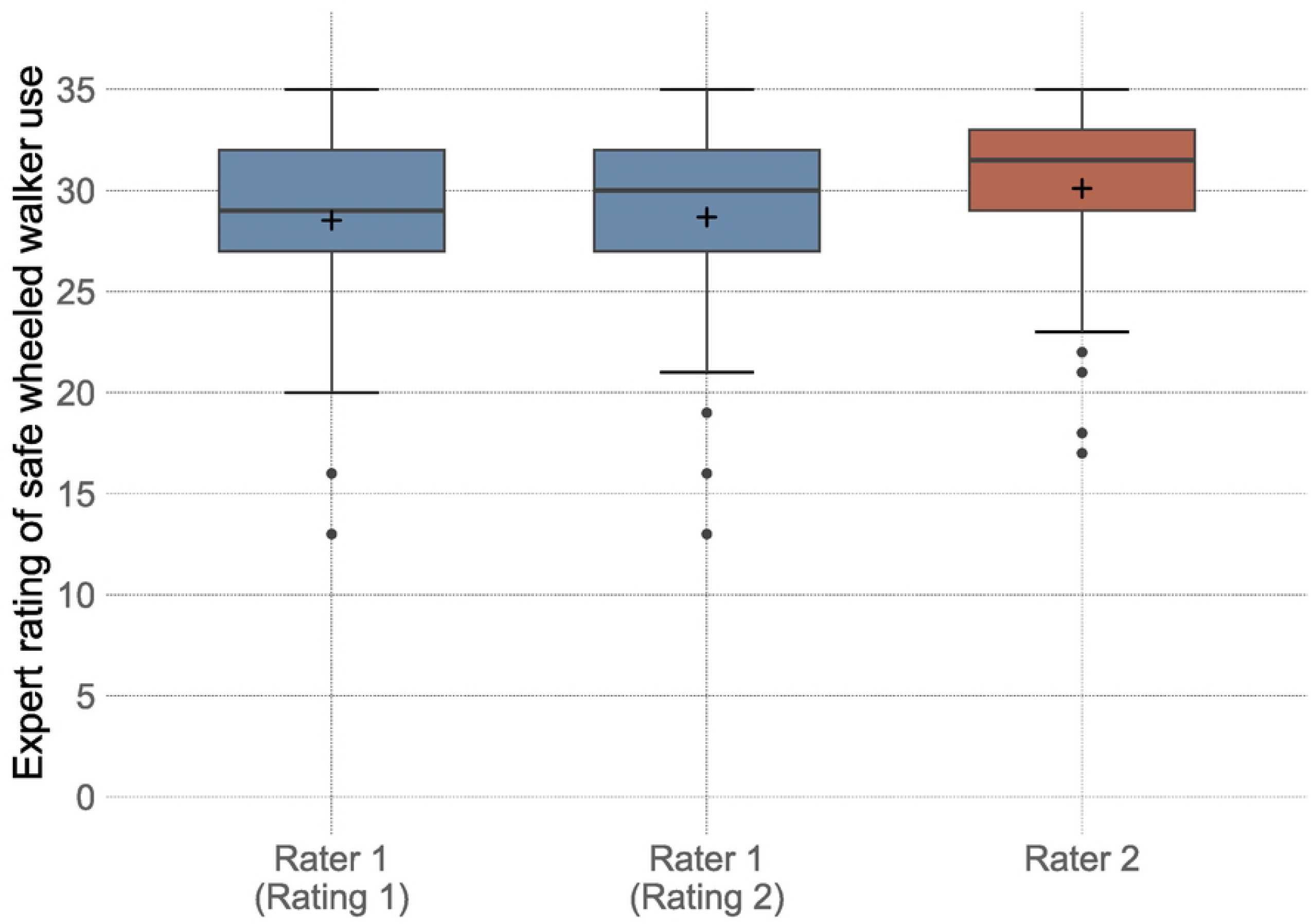
Boxplot with mean values (+) of the expert ratings of safe wheeled walker use (score range 0-35, higher values indicate maximum safety) for rater 1 (rating 1: first rating, rating 2: second rating after 7 days) and rater 2

Results of reliability testing were ICC_3.1_ = 0.98 for intra-rater reliability and ICC_2.1_ = 0.80 for inter-rater reliability (**Table 3**). Validity testing yielded correlation coefficients ranging from r = -0.45 for comparison between the expert rating by Rater 1 and the Timed Up and Go test to r = 0.74 for the comparison between the expert rating by Rater 1 and nursing staff ratings. Differences in correlation coefficients between the two raters were small with r = 0.1 or less.

Bland–Altman analyses indicated that participants and nursing staff rated safety lower than experts, with mean differences of 3.23 points (95% limits of agreement: -7.38 to 13.84) and 5.63 points (95% limits of agreement: -5.50 to 16.76), respectively (**Figure 3**). After two weeks, clinic patients (n = 29) showed a mean improvement of 2.38 points, corresponding to 8.4% of the baseline mean score. Reasons for non-participation in the follow-up assessment (n=11) included premature discharge and transfer to the acute care unit.

**Table 3.** Reliability (n = 50), validity (n = 50), and sensitivity to change (n = 29) of the expert rating in 40 clinic patients and 10 nursing home residents.

|  |  |
| --- | --- |
| Intra-rater reliability Rater 1, R1 vs. R2, ICC<sub>3,1</sub> | 0.98 (95%-CI: 0.96; 0.99) |
| Inter-rater reliability Rater 1 R1 vs. Rater 2 R1, ICC<sub>(2,1)</sub> | 0.80 (95% CI: 0.53; 0.90) |
| Rater 1 R1, expert rating vs. Timed Up and Go test, r | -0.45 (95% CI: -0.64; -0.19) |
| Rater 2 R1, expert rating vs. Timed Up and Go test, r | -0.49 (95% CI: -0.67; -0.24) |
| Rater 1 R1, expert rating vs. self-rating, r | 0.50 (95% CI: 0.25; 0.68) |
| Rater 2 R1, expert rating vs. self-rating, r | 0.40 (95% CI: 0.14; 0.61) |
| Rater 1 R1, expert rating vs. nursing staff rating, r | 0.74 (95% CI: 0.59; 0.85) |
| Rater 2 R1, expert rating vs. nursing staff rating, r | 0.73 (95% CI: 0.56; 0.84) |
| Sensitivity to change Rater 1 R1 vs. R3 (n = 29) |  |
| Mean difference | 2.38 (95% CI: 1.22; 3.54) |
| Cohen's D | 0.58 (95% CI: 0.28; 0.88) |
| Standardized Response Mean (SRM) | 0.78 (95% CI: 0.47; 1.25) |

R1 = rating 1; R2 = rating after 7 days; R3 = assessment and rating 2 weeks later; ICC = intra-class coefficients of correlation; 95%-CI = 95% confidence interval; r = Person's coefficient of correlation

**Figure 3.**
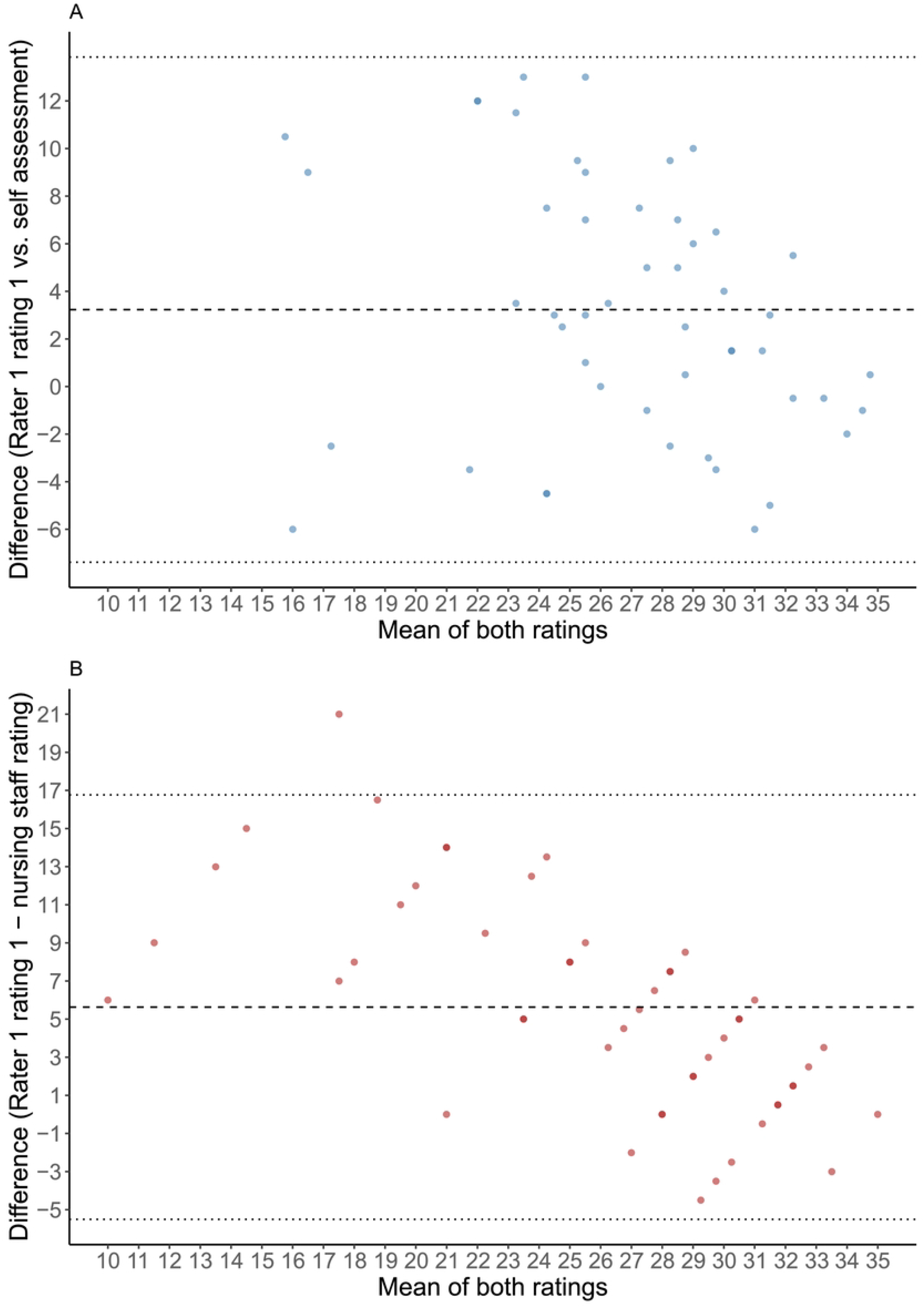
Bland–Altman plots comparing Rater 1 with self-assessment (A) and nursing staff assessment (B). Each point represents one participant. Dashed horizontal lines indicates the mean difference (bias), and the dotted lines represent the 95% limits of agreement (mean difference ± 1.96 SD).

## Discussion

This study aimed to develop and evaluate a video-based expert rating instrument for assessing safe wheeled walker use in frail older adults. During the initial focus group meetings, it became apparent that, given the five criteria to be assessed simultaneously for each hazardous situation, video recording followed by rating was necessary. The findings demonstrated excellent intra-rater reliability, good inter-rater reliability, and acceptable validity when compared with the TUG test, nursing staff ratings, and participants’ self-assessments. As no gold standard exists for assessing safe wheeled walker use in frail older adults, convergent validity was used for validity evaluation. In addition, the instrument was sensitive to change, detecting improvements in safe wheeled walker use after a two-week rehabilitation period.

Although the study cohort was heterogeneous, all participants showed functional impairments as indicated by TUG performance exceeding 15 seconds, even accounting for the possible time delay associated with walker use [15]. This underscores the relevance of the instrument for frail populations in geriatric rehabilitation and long-term care settings.

The excellent intra-rater reliability and good inter-rater reliability indicate that the rating system can be applied consistently over time and across different raters. These findings are in line with reports from comparable observational instruments in geriatrics [11, 12]. The strong correlation with nursing staff ratings supports convergent validity, whereas the moderate negative correlation with the TUG test suggests that physical performance and safe wheeled walker use are related but distinct constructs. This distinction is clinically important, as safe wheeled walker use depends not only on physical capacity but also on appropriate handling of the device in potentially hazardous situations, which are not fully captured by the TUG test [13].

Although the assessment could be conducted and rated successfully, the time required in its current form may limit routine clinical application. This highlights the need for a shorter version focusing on the most discriminative tasks to enhance feasibility without compromising reliability and validity. Such a streamlined instrument would be particularly valuable in geriatric rehabilitation clinics and nursing homes, where time and staffing resources are limited. Analysis of frequently occurring negative safety criteria suggests that brake handling, turning on the spot (180°), sitting down on the walker, and opening a door are particularly relevant for safety. A shorter version focusing on these high-risk situations could substantially reduce assessment time while still capturing the most essential aspects of safe wheeled walker use. In its current form, the instrument remains suitable for research or for more detailed individual assessment in clinical and nursing home settings.

Digital solutions, including tablet-based applications and artificial intelligence (AI) supported video analysis, may further enhance efficiency, scalability, and objectivity. Automated detection of unsafe movement patterns could reduce rater dependency, enable real-time feedback, and facilitate broader implementation in both clinical and home-based settings.

Compared with the Safe Use of Mobility Aid Checklist [9], the present instrument offers several advantages for frail, institutionalized older adults. Hunter’s checklist was designed for individuals with dementia and includes outdoor tasks that are not always feasible in hospitals or nursing homes. By focusing exclusively on indoor scenarios, the present instrument is better suited to clinical and residential care environments, where fall risk is high and standardized testing conditions are needed.

The demonstrated sensitivity to change, reflected by measurable improvements after two weeks of standard rehabilitation, suggests that the instrument may serve purposes beyond basic risk assessment. Specifically, it may be useful for monitoring therapeutic effects, guiding individualized interventions, and documenting progress, all of which are important in multifactorial fall prevention programs [14].

Several limitations should be acknowledged. First, the study sample was relatively small and geographically restricted, which may limit generalizability. Second, although video-based assessments enhance standardization, it may not fully capture contextual factors present during live assessments, such as environmental distractions or therapist–patient interactions. Finally, while inter-rater reliability was good, it did not reach the excellent range, suggesting that further refinement of rating instructions or standardized rater training could improve consistency.

## Conclusions

In conclusion, this study presents the development and validation of a novel video-based expert rating instrument for assessing safe wheeled walker use in frail older adults. The instrument demonstrated high reliability, acceptable validity, and sensitivity to change. In its current version, it is suitable for research or in detailed individual assessment in clinical and nursing home settings, but may be too time-consuming for routine clinical use. With further refinements, particularly through the development of a shorter version and the integration of AI- supported rating, the instrument has the potential to become a widely used tool for identifying unsafe wheeled walker use, tailoring and evaluating interventions, and reducing fall risk in vulnerable populations.

## Data Availability

The minimal data set is available upon request from the corresponding author.

## Acknowledgment

Special thanks goes to physiotherapists Melanie Kallmes and Daniela Arndt for their valuable assistance with the video rating.

